# Cholesteryl Ester Transfer Protein as a Drug Target for Cardiovascular Disease

**DOI:** 10.1101/2020.09.07.20189571

**Authors:** Amand F Schmidt, Nicholas B Hunt, Maria Gordillo-Marañón, Pimphen Charoen, Fotios Drenos, Juan-Pablo Casas, Mika Kivimaki, Deborah A Lawlor, Claudia Giambartolomei, Olia Papacosta, Nishi Chaturvedi, Joshua C Bis, UCLEB consortium, Christopher O’Donnell, Goya Wannamethee, Andrew Wong, Jackie F Price, Alun D Hughes, Tom R Gaunt, Nora Franceschini, Dennis O Mook-Kanamori, Magdalena Zwierzyna, Reecha Sofat, Aroon D Hingorani, Chris Finan

**Author notes:** Shared senior authors (Aroon D Hingorani, Chris Finan). **Corresponding author** Amand Floriaan Schmidt, PhD.

## Abstract

Drug development of cholesteryl ester transfer protein (CETP) inhibition to prevent coronary heart disease (CHD) has yet to deliver licensed medicines. To distinguish compound from drug target failure, we compared evidence from clinical trials and Mendelian randomization (MR) results. Findings from meta-analyses of CETP inhibitor trials (≥ 24 weeks follow-up) were used to judge between-compound heterogeneity in treatment effects. Genetic data were extracted on 190+ pharmacologically relevant outcomes; spanning 480,698-21,770 samples and 74,124-4,373 events. Drug target MR of protein concentration was used to determine the on-target effects of CETP inhibition and compared to that of PCSK9 modulation. Fifteen eligible CETP inhibitor trials of four compounds were identified, enrolling 79,961 participants. There was a high degree of heterogeneity in effects on lipids, lipoproteins, blood pressure, and clinical events. For example, dalcetrapib and evacetrapib showed a neutral effect, torcetrapib increased, and anacetrapib decreased cardiovascular disease (CVD); heterogeneity p-value < 0.001. In drug target MR analysis, lower CETP concentration (per μg/ml) was associated with CHD (odds ratio 0.95; 95%CI 0.91; 0.99), heart failure (0.95; 95%CI 0.92; 0.99), chronic kidney disease (0.94 95%CI 0.91; 0.98), and age-related macular degeneration (1.69; 95%CI 1.44; 1.99). Lower PCSK9 concentration was associated with a lower risk of CHD, heart failure, atrial fibrillation and stroke, and increased risk of Alzheimer’s disease and asthma. In conclusion, previous failures of CETP inhibitors are likely compound related. CETP inhibition is expected to reduce risk of CHD, heart failure, and kidney disease, but potentially increase risk of age-related macular disease.

## Introduction

The causal role of low-density lipoprotein cholesterol (LDL-C) in coronary heart disease (CHD) has been established through randomized controlled trials (RCTs) of different LDL-C lowering drug classes^1 2 3,4^ and by Mendelian randomization (MR) studies^5^.

Circulating high-density lipoprotein cholesterol (HDL-C) shows an inverse association with CHD in nonrandomized studies^6^. MR studies utilizing genetic variants associated with HDLC selected throughout the genome have provided inconclusive evidence on the causal role of HDL-C as a biomarker^5^’^7^. Findings from RCTs of niacin^8^ and cholesteryl ester transfer protein (CETP) inhibitors^9^, developed to prevent CHD by raising HDL-C have also been disappointing. For example, of the four CETP inhibitors that have progressed to phase 3 clinical trials, none have received market authorization (Supplementary Table 1). Six other CETP inhibitors (e.g. obicetrapib) are still in active development, raising important questions about the validity of CETP as a therapeutic target^10^. One interpretation is that HDL-C is not causally related to CHD, and that raising HDL-C as a therapeutic strategy will be an ineffective approach for CHD prevention. As a result, the reduction in CHD events observed in a large RCT of anacetrapib (odds ratio [OR] 0.91 95%CI 0.85; 0.97)^11^, was attributed to its effect on LDL-C rather than to its HDL-C raising action^10^.

However, analysis of lipoprotein sub-classes measured using nuclear magnetic resonance (NMR) spectroscopy suggests that, unlike LDL-C, HDL-C particles encompasses several lipoprotein sub-fractions that have differential associations with CHD: some fractions being associated with higher and others with lower CHD risk ^12,13^. Additionally, failures of CETP inhibitors might be related to the developed compounds rather than the drug target itself, either because of inadequate target engagement or a competing off-target action.

Compound related failures can be addressed by developing an improved CETP inhibitor, whereas target failure affects *all* CETP inhibitors.

To address these uncertainties, we performed a drug target MR study^14^ of CETP, focusing on variants within the encoding gene (acting in *cis)* that are associated with circulating CETP concentration, to directly model the effects of pharmacological action on this target by a clean drug with no off-target actions. To evaluate potentially diverse effects of drug target perturbation, we combined drug target MR with a phenome-wide scan of over 190 disease biomarkers or clinical end-points relevant to cardiovascular as well as non-cardiovascular outcomes^15^. We compared drug target MR effect estimates to compound-specific effect estimates derived from a systematic review and meta-analysis of CETP inhibitor RCTs. Assuming the developed CETP inhibitors sufficiently engaged the drug target, on-target failures would result in consistent treatment effects across all compounds, which should be similar to the on-target effect modelled through MR. Finally, drug target MR analyses of CETP and PCSK9, an archetypal LDL-C lowering drug target, were compared on their effects profile.

## Methods

### Systematic review and meta-analyses of CETP inhibitor effects

CETP inhibitor trials with at least 24 weeks of follow-up (irrespective of phase) were identified through a systematic review using a pre-specified search strategy (See Appendix) of MEDLINE and OVID, supplemented by clinicaltrials.gov. Parallel-group RCTs were included regardless of comparator (placebo or active therapy) with no additional exclusion criteria. Treatment effects were extracted (by NH and AFS) on lipids, lipoproteins, blood pressure, the incidence of all-cause mortality (ACM) and cardiovascular endpoints: any cardiovascular disease (CVD, defined as CV death, myocardial infarction (MI), any stroke, and angina hospitalization), fatal CVD (FCVD), any MI (including CHD), fatal MI (FMI), any stroke (ST; including ischemic, hemorrhagic and other strokes), ischemic stroke (IST), hemorrhagic stroke (HST), and heart failure (HF). Treatment effects on continuous traits (mean differences) were extracted as the between group difference in change from baseline^16^. Additional data were extracted on compound dose and potency, trial participants, and setting. Compound-specific clinical trial data were meta-analysed using the inverse-variance weighted method using both fixed and random effects. We used the Q-statistic^17,18^ to test for the presence of between compound heterogeneity.

### Mendelian randomisation analysis

Drug target MR analysis^14^ utilises (cis)-variants in, or near, a drug target encoding gene to obtain a causal estimate of the protein effect on multiple outcomes. Specifically, genetic associations with an outcome (e.g. CHD) are regressed on genetic associations with the drug target protein concentration or, alternatively, with biomarkers distal to the protein.

Under the assumption that all the effects of the genetic variants on an outcome are mediated by the drug target protein (no-horizontal pleiotropy), the slope represents an estimate of the drug target effect. Here we used genetic effect estimates on the concentration of the encoded protein (CETP or PCSK9) as the primary exposure of interest, repeating the analyses using genetic effect association with LDL-C (for CETP and PCSK9), HDL-C (for CETP), and triglycerides (TG; for CETP), representing biomarkers known to be affected by the corresponding protein (available from the GLGC^19^ consortium).

To reduce the risk of “weak-instrument bias”^20^, we selected genetic variants with an F-statistic of 15 or higher (Supplemental Tables 2-7). We used a two-staged MR-paradigm, where genetic associations with the exposure and outcome were derived in independent samples, ensuring that any remaining weak-instrument bias attenuates towards the null (conservative estimates)^20^. Given the differences in coverage between the various outcome GWAS, variants were clumped to an R-squared of 0.40 *after* linking the exposure variants to a specific outcome GWAS (maximizing precision). Residual linkage disequilibrium (LD) was modelled using a generalised least squares (GLS)^21,22^ IVW-estimator, and an external correlation structure (random 5,000 UK biobank, (UKB) sample). The possibility of bias inducing horizontal pleiotropy was minimized by focussing on a *cis* genetic region, excluding variants with large leverage or outlier statistics^14,23^ and using the Q-statistic to identify remaining violations^23,24^.

Findings from the *cis-MR* analysis of CETP were compared to effects observed in trials (for outcomes shared by the trial and MR analyses) using hierarchical clustering.

### Selection of genetic instruments

Genetic associations with CETP concentration (protein quantitative trait loci; pQTLs) were extracted from a GWAS on circulating CETP concentration^25^. Genetic variants were selected based on residency within a narrow window around *CETP* (Chr 16: bp: 56,961,923 to 56,985,845; GRCh38)^25^. For the PCSK9 drug target MR, we selected variants associated with PCSK9 concentration^26^ using the following window: 55,037,447 to 55,066,852 bp (Chr 1; GRCh38). *CETP* variants with a minor allele frequency (MAF) below 0.05 were removed. For *PCSK9*, this threshold was reduced to 0.01, ensuring rs11591147 was included (the top hit with PCSK9 concentration^26^).

### Genetic associations with outcomes of interest

GWAS data were available for over 190 outcome traits (see Supplementary Methods and Table 8) including 60,801 CHD cases from CardiogramplusC4D^27^; 40,585 stroke cases (subtypes) from MEGASTROKE^28^; 47,309 HF cases from HERMES^29^, 60,620 atrial fibrillation (AF) cases from AFgen ^30^, 71,880 Alzheimer’s disease (AD)^31^ cases from a metaanalysis of the PGC-ALZ, IGAP, 16,144 age-related macular degeneration (AMD) events from IAMDGC^32 33^, and genetic associations with NMR measured circulating lipoprotein subfractions and other metabolites were available from a meta-analysis of Kettunen *et al^34^*, and UCLEB^35^ (n: 33,029).

Results are presented as mean difference (MD) or odds ratio (OR) with 95% confidence interval (95%CI) coded towards the drug target effect direction; i.e., towards lower circulating protein, LDL-C, and TG concentration, and a higher HDL-C concentration. CETP concentration was reported as g/ml while PCSK9 concentration was reported as log-transformed ng/ml.

## Results

### Effects of different CETP inhibitors in trials

We identified 15 RCTs of CETP inhibitors with at least 24 weeks of follow-up, including four different compounds (six anacetrapib, four dalcetrapib, four torcetrapib and one evacetrapib study), all evaluated against placebo (Supplementary Table 9) and involving 79,961 participants. Participants received either torcetrapib 60-120 mg, evacetrapib 130 mg, anacetrapib 100mg, or dalcetrapib 600-900 mg per day, reflecting differences in compound potency (Supplementary Table 9, and Supplementary results). The longest follow-up times was a median of 49 months for anacetrapib in the REVEAL trial, 31 months for dalcetrapib in the DAL-OUTCOMES trial, 24 months for torcetrapib in the RADIANCE 1 and ILLUSTRATE trials and 26 months for evacetrapib ACCELERATE trial.

All four compounds increased HDL-C and reduced LDL-C, but the magnitude of effect differed between compounds (Figure 1, Supplement Table 10). Anacetrapib and evacetrapib had the largest HDL-C increasing effect, 130% (95%CI 127; 133) and 132% (95%CI 130; 133) respectively, followed by torcetrapib 52% (95%CI 49; 55) and dalcetrapib 29% (95%CI 23; 43); heterogeneity p-value < 0.001. The reduction in LDL-C was -38% (95%CI -40; -36) for anacetrapib, -37% (95%CI -38; -36) for evacetrapib, -20% (95%CI -24; -17) for torcetrapib, and -1% (95%CI -1.1; -0.9) for dalcetrapib. The CETP inhibitor effects were similarly heterogenous (interaction p-value < 0.001) for TG, apolipoprotein A1, B, lp(a), and systolic/diastolic blood pressure (SBP/DBP); Figure 1 and Supplemental Figure 1 and Table 10.

**Figure 1.**
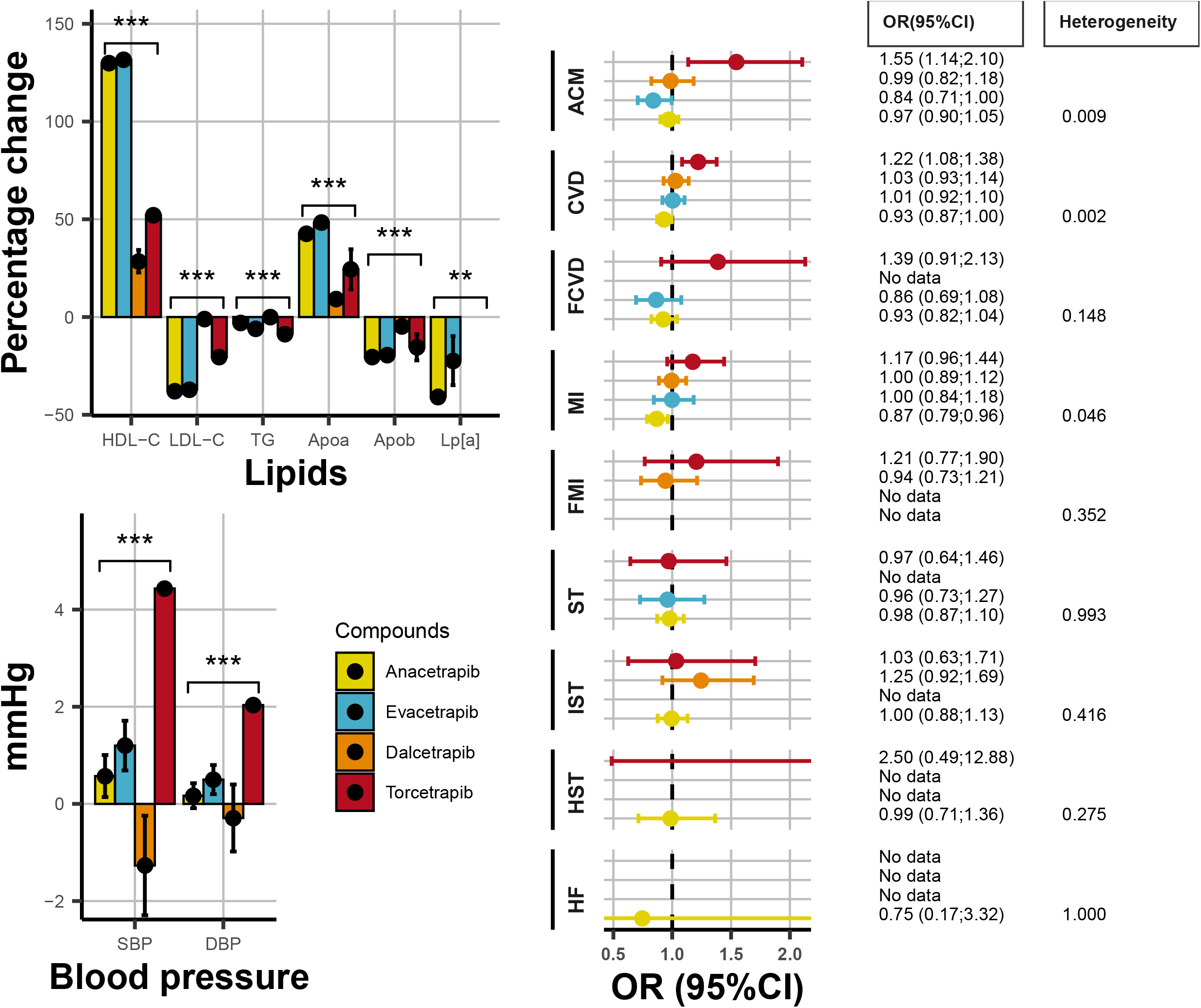
Differences in CETP-inhibitor effects on lipids, blood pressure and clinical endpoints. N.B Results are based on a fixed effect compound specific meta-analyses with differences between compounds tested using a Q-test (Heterogeneity). *** indicates a p-value < 0.001 for the Q-test. LDL: LDL-C, HDL: HDL-C, TG: triglycerides, ApoA1: apolipoprotein A1, ApoB: apolipoprotein B, S/DBP: systolic/diastolic blood pressure ACM: All-cause mortality, CVD: cardiovascular disease, FCVD: fatal-CVD, MI: myocardial infarction, FMI: fatal-MI, ST: any stroke, IST: Ischemic stroke, HST: haemorrhagic stroke, HF: heart failure.

CETP inhibitors also differed in their effect on clinical outcomes (Figure 1). Torcetrapib increased risk of all-cause mortality (OR 1.56 95%CI 1.14; 2.12), while evacetrapib decreased all-cause mortality (OR 0.84 95%CI 0.71; 1.00); heterogeneity p-value = 0.009. Similarly, torcetrapib increased any CVD (OR 1.22 95% 1.08; 1.38), while anacetrapib decreased CVD (OR 0.93 95%CI 0.87; 1.00); heterogeneity p-value 0.002. Anacetrapib reduced any MI risk (OR 0.89 95%CI 0.80; 0.99), with the remaining compounds showing a neutral MI effect; heterogeneity p-value 0.046.

### On-target effects of CETP inhibition using drug target MR

Lower genetically instrumented CETP concentration was associated with lower LDL-C -0.08 (mmol/L, 95%CI -0.08; -0.08), TG -0.09 (mmol/L, 95%CI -0.09; -0.09), Lp[a] -2.13 (nmol/L, 95%CI -1.52; -2.74), apolipoprotein B -0.03 (g/L, 95%CI -0.03; -0.03), and higher HDL-C 0.24 (mmol/L, 95%CI 0.24; 0.24), and apolipoprotein A1 0.14 (g/L, 95%CI 0.14; 0.14); Figure 2 (full details in Supplementary Table 11).

**Figure 2.**
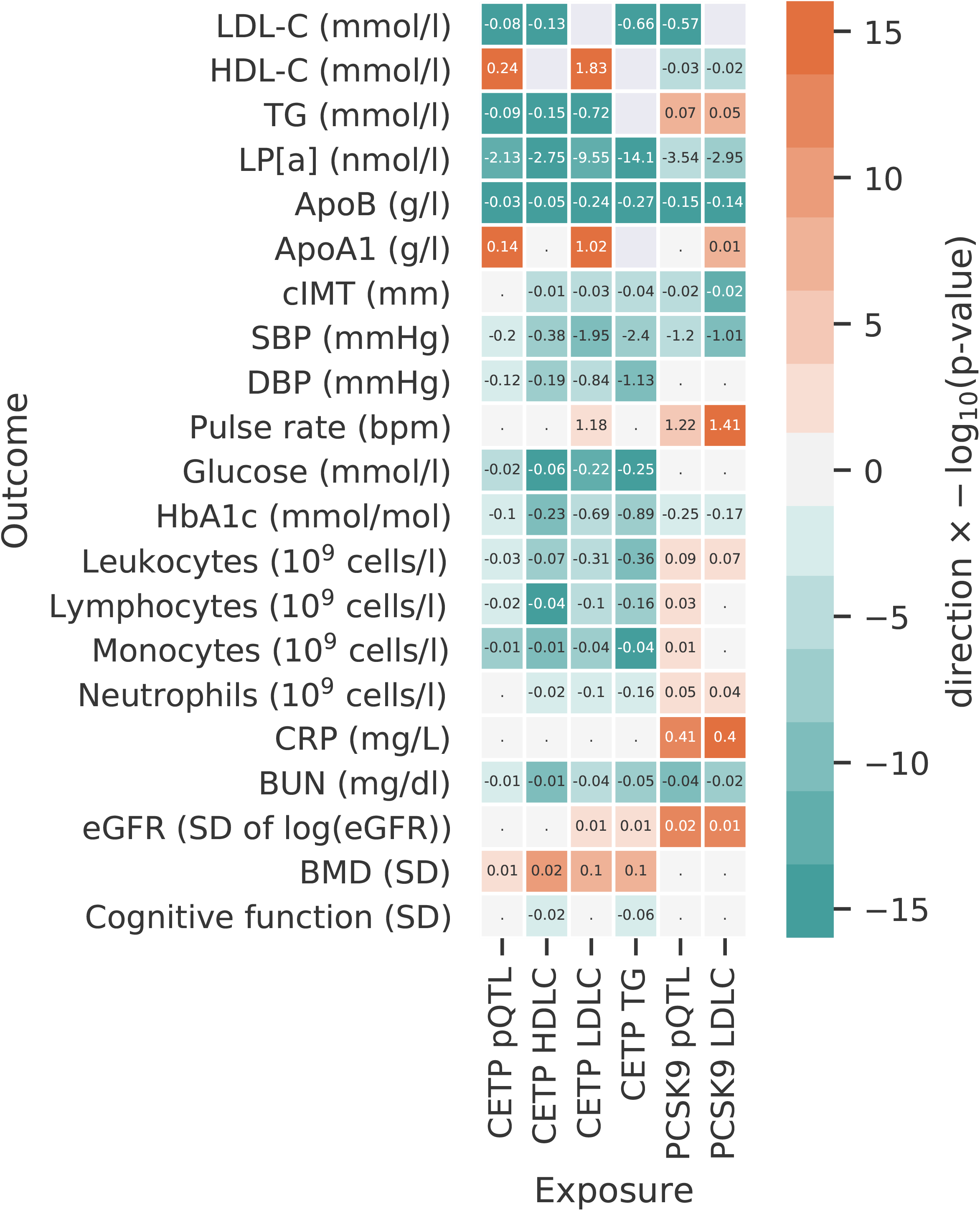
Drug target Mendelian randomization estimates of lower CETP and PCSK9 weighted by genetic associations with protein concentration or downstream lipid. N.B The rows represent the quantitative outcomes and the columns represent the intermediate variables (approximating) drug target concentration. Cells are coloured by effect direction times -log_10_(p-value), with the mean difference (the slope coefficient) provide for MR results with a p-value smaller than 0.05. The p-values was truncated at 10^-16^ ensuring sufficient variation in the colour code.

Lower CETP concentrations were significantly associated with a lower blood pressure (−0.2 mmHg for SBP and -0.12 for DBP), lower concentration of blood glucose (−0.02 mmol/L), HbA1c (−0.10 mmol/mol), lower cell counts for leukocytes (−0.03 x 10^9^ cells/L), lymphocytes (−0.02 x 10^9^ cells/L), and monocytes (−0.01 x 10^9^ cells/L). These findings were consistent in cis-MR analysis weighted by LDL-C, HDL-C and TG (Supplemental Figure 2).

Lower genetically instrumented CETP concentration was associated with CHD (OR 0.95; 95%CI 0.91; 0.99), HF (OR 0.95; 95%CI 0.92; 0.99), CKD (OR 0.94 95%CI 0.91; 0.98), and AMD (OR 1.31 95%CI 1.22; 1.40); Figure 3 and Supplemental Table 12. The magnitude and direction of effects were consistent in LDL-C, HDL-C and TG weighted analyses (Supplemental Figure 2, Table 12).

**Figure 3.**
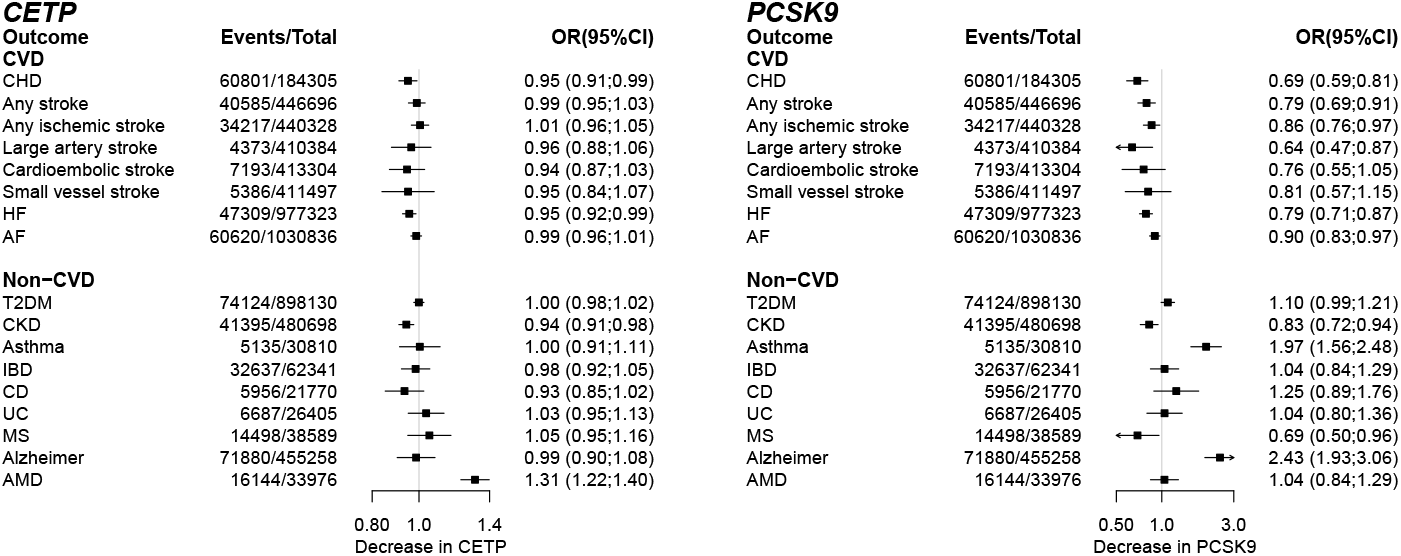
The drug target Mendelian randomization effects of lower CETP and PCSK9 concentration on clinical end-points. N.B CHD: coronary heart disease, HF: heart failure, AF: atrial fibrillation, T2DM: type 2 diabetes mellitus, CKD: chronic kidney disease, IBD: inflammatory bowel disease, CD: Crohn’s disease, UC: ulcerative colitis, MS: multiple sclerosis, AMD: age-related macular degeneration.

### On-target effects of PCSK9 inhibition using drug target MR

We compared the drug target MR results of CETP lowering to those for PCSK9, using genetic instruments on PCSK9 concentration. Lower PCSK9 concentration (Figure 2, Supplemental Table 11) was associated with lower LDL-C (−0.57 mmol/L), apolipoprotein B (−0.15 mmol/L), lp[a] (−3.54 nmol/L) and HDL-C (−0.03 mmol/L). We additionally observed an association with carotid intima-media thickness (−0.02 mm), SBP (−1.20 mmHg), blood urea nitrogen (BUN: -0.04 mg/dl), HbA1c (−0.25 mmol/mol), and higher estimated-GFR (eGFR: 0.01 per SD), C-reactive protein (CRP: 0.41 mg/L), pulse rate (1.22 bpm), and blood cell counts (Figure 2, and Supplemental Table 11).

Lower PCSK9 concentration was associated with the following clinical endpoints (Figure 3, Supplementary table 12): CHD (OR 0.69 95%CI 0.59; 0.81), any stroke (OR 0.79 95%CI 0.69; 0.91), any ischemic stroke (OR 0.86 95%CI 0.76; 0.97), large artery stroke (OR 0.64 95%CI 0.47; 0.87), HF (OR 0.79 95%CI 0.71; 0.87), AF (OR 0.90 95%CI 0.83; 0.97), CKD (OR 0.83 95%CI 0.72; 0.94), multiple sclerosis (MS; OR 0.69 95%CI 0.50; 0.96), and increased risk of asthma (OR 1.97 95%CI 1.56; 2.48) and AD (OR 2.43 95%CI 1.93; 3.06). The LDL-C weighted analysis was consistent with these findings (Figure 2, Supplemental Figure 2 and Table 12).

### Lipoprotein sub fraction profiles based on NMR spectroscopy

Drug target MR showed that lower CETP concentration was associated with wide ranging effects on lipoprotein sub-fraction size and content including medium, large and extra-large HDL-C subfractions, and lower extra-small, small, and medium VLDL sub-fractions (Figure 4). Lower CETP concentration had a minimal effect on total LDL-C measured through NMR spectroscopy: -0.01 SD (95%CI -0.05; 0.02) for LDL-C compared to a HDL-C effect of 0.51 SD (95%CI 0.46; 0.56). Lower CETP was however strongly associated with decreased mean LDL-C diameter -0.24 SD (95%CI -0.29; -0.20). The PCSK9 NMR profile was narrower than that for CETP, with lower PCSK9 associated only with lower medium and large LDL-C subfractions, IDL, and extra small VLDL.

**Figure 4.**
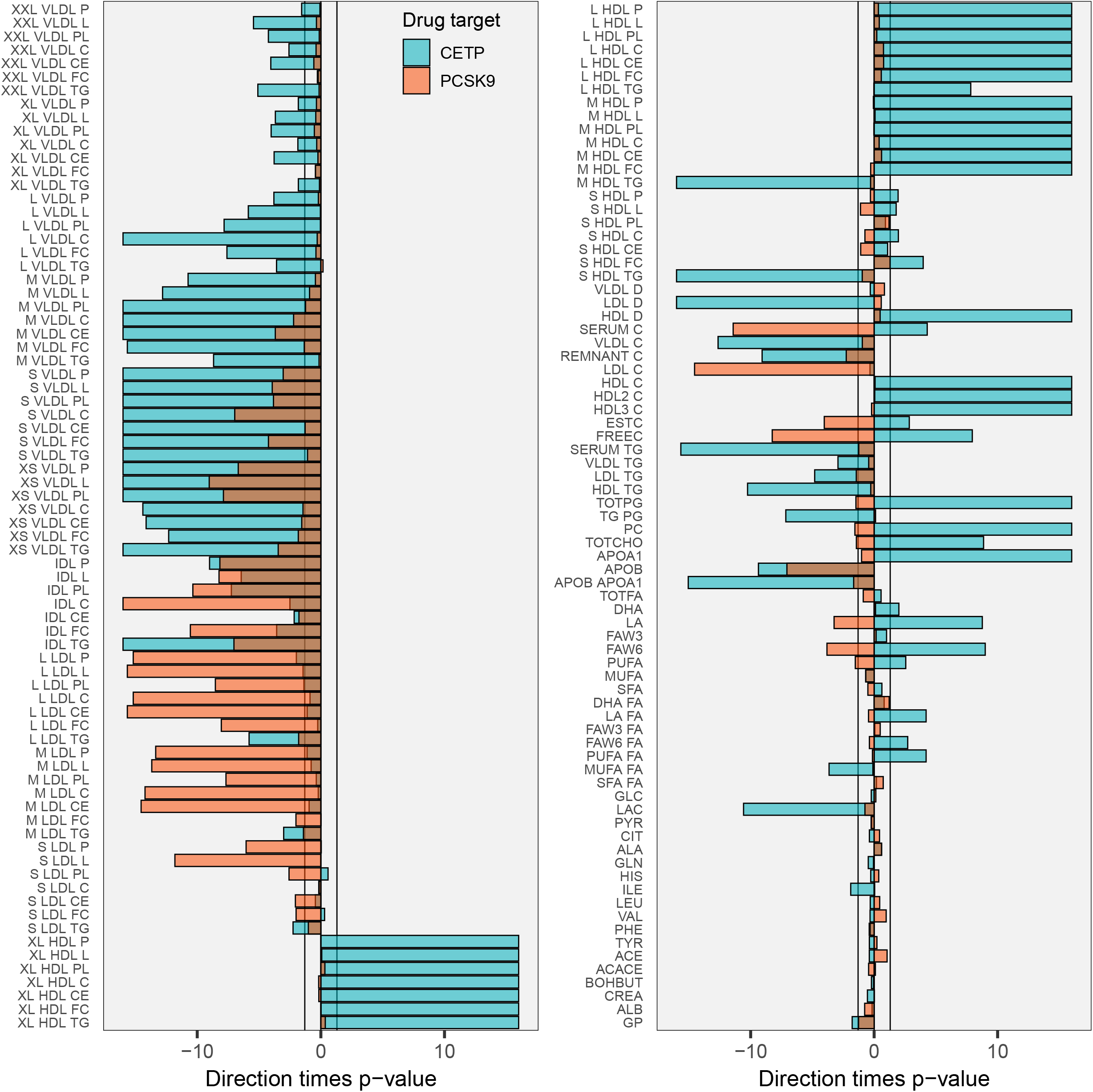
The drug target Mendelian randomization effects of lower CETP and PCSK9 concentration on NMR-measured metabolites. N.B Results are provided as -log_10_(p-values) times effect direction, with the x-axis limits set to ±16. Bars semi-transparent and plotted on-top of each other to directly compare the two drug targets in their NMR measured lipids effect estimates. The vertical lines at ±1.3 represent the traditional p-value threshold of 0.05.

### Comparing effects of CETP inhibitors to drug target MR effects of CETP modulation

CETP inhibitors could be directly compared to the on-target MR effects of lower CETP concentration for their effect on lipids, lipoprotein, blood pressure and any MI (Figure 5).

**Figure 5.**
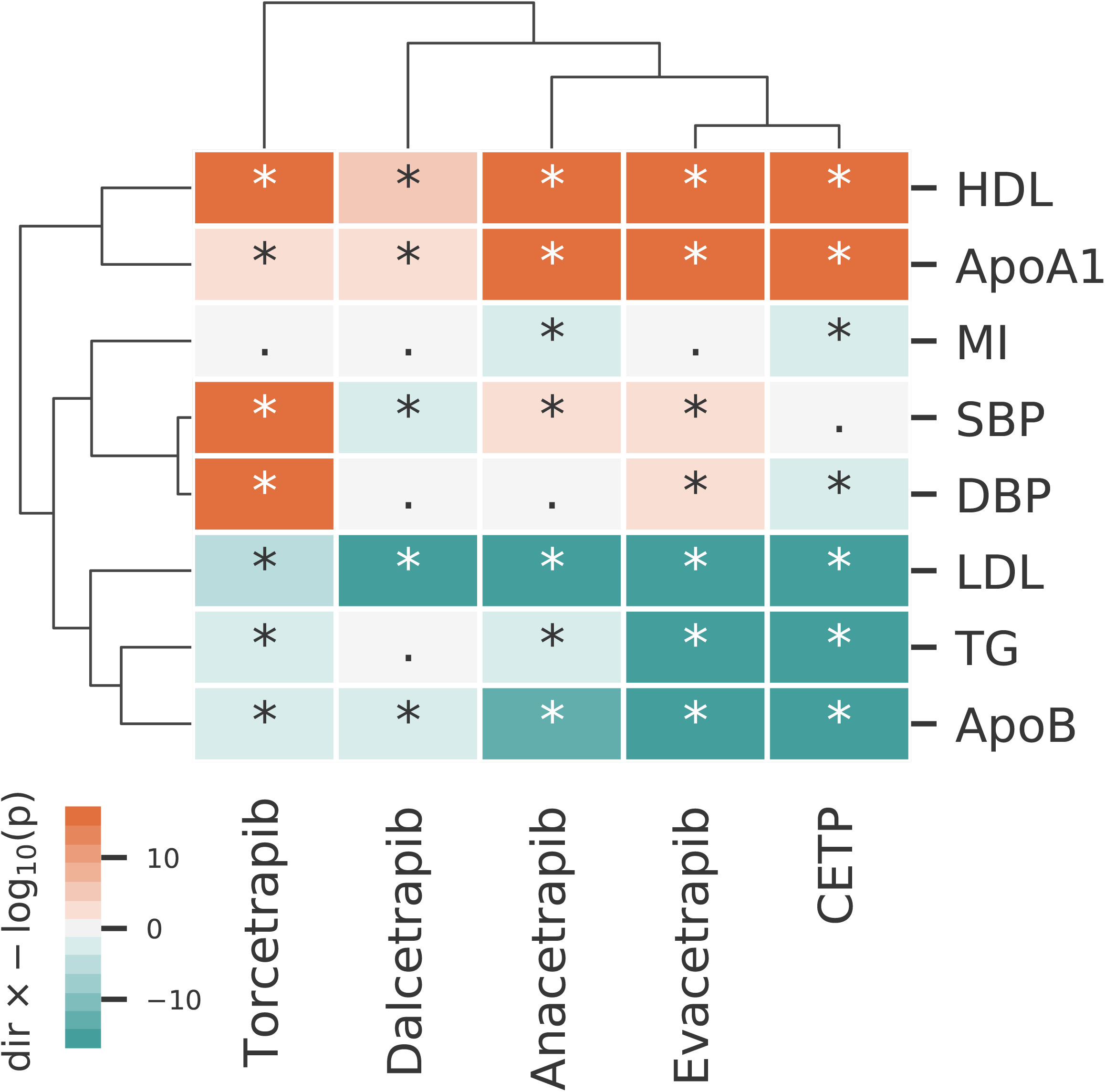
A cluster analysis comparing the on-target Mendelian randomization effect of lower CETP concentration to effects from CETP inhibiting compounds. N.B Clustering was performed on the square root of the -log_10_(p-values) x effect direction, with the p-value truncated to 10^-60^ to ensure enough difference between the CETP compound effect on changes in lipids. Associations with a p-value below 0.05 are indicated with a star. The dendrograms represent clustering by outcome (rows) and compound/drug target (columns).

Both torcetrapib and dalcetrapib showed biomarker profiles distinct from that of genetically instrumented lower CETP concentration. For torcetrapib this difference was driven by an increasing effect on SBP and DBP. For dalcetrapib this difference was due to attenuated lipid associations. Anacetrapib and evacetrapib displayed a similar risk factor profile that most closely reflected the on-target association of lower CETP concentration modelled genetically, and hence clustered most closely to on-target CETP modulation.

## Discussion

We found substantial heterogeneity in the effects of four CETP-inhibitors (anacetrapib, evacetrapib, dalcetrapib and torcetrapib) on major lipid fractions, blood pressure, all-cause mortality and cardiovascular outcomes, suggesting between-compound differences in the efficacy of CETP inhibition, off-target actions or both. The effects profile of anacetrapib and evacetrapib on blood lipids and cardiovascular end-points most closely matched the effects of genetically-instrumented reductions in CETP concentration suggesting that anacetrapib and evacetrapib are effective CETP inhibitors.

The reduction in cardiovascular events seen in the REVEAL trial of anacetrapib (median follow-up 1,497 days; Supplementary Table 3) is consistent with the drug target MR results presented here. The manufacturer, Merck, did not seek marketing authorization for this drug citing an anticipated lack of regulatory support^36^. The evacetrapib ACCELERATE trial was terminated for futility after a median follow-up of 791 days, a time point before the benefits of anacetrapib emerged in the REVEAL trial (see Figure 1 of ref^11^). Taken together, the presented RCT and drug target MR findings, suggest that CETP is a viable target to manage CVD risk. The heterogeneous clinical effects of evaluated CETP inhibitors, e.g. the increased risk of mortality and CVD by torcetrapib or the modest LDL-C effect of dalcetrapib, are likely to be compound - rather than target-related ^37^.

As well as enabling a separation of on-vs off-target effects of CETP inhibition, drug target MR analysis facilitate an investigation of CETP effect beyond those investigated in clinical trials. The drug target MR analyses showed that lower CETP concentration was additionally associated with not only with CHD (OR 0.95 per µg/ml CETP concentration; 95%CI 0.91; 0.99), but also HF (OR 0.95; 95%CI 0.92; 0.99) and CKD (OR 0.94; 95%CI 0.91; 0.98), but with a higher risk of AMD (OR 1.31; 95%CI 1.22; 1.40). Similar to the on-target effects of CETP, genetically-instrumented PCSK9 concentration was associated with a lower risk of CHD, HF and CKD, and additionally with any stroke, ischemic stroke, AF, MS, as well as an increased risk of asthma and AD^38^. We showed that CETP and PCSK9 had distinct effect patterns on different lipoprotein sub-fractions, with lower CETP being associated with higher HDL-C and lower VLDL-C sub-fractions, and PCSK9 with lower LDL-C sub-fractions alone. These findings suggest that, although sharing salutary effects on clinical endpoints, the mechanisms through which the effects of CETP and PCSK9 inhibition are mediated are likely to be target-specific and cannot, on present evidence, be attributed to selected shared actions or a single pathway e.g. on LDL-C or apolipoprotein B^39^.

Some prior drug target MR studies have attempted to quantify the anticipated effect of a drug targeting the same protein. For example, the anticipated effect of CETP inhibition on CHD risk is a reduction of 40% when weighted by one mmol/L lower LDL-C concentration (Supplemental figure S2). While of potential interest, there are some caveats that suggest that drug target MR analysis may be more useful as a reliable test of effect direction, and when multiple outcomes are considered, the rank order of effects. This is because drugs that inhibit a target do so usually by modifying its function not its concentration, whereas genetic variants used in MR analysis usually affect protein expression and therefore concentration. However, for enzymes like CETP, activity reflects both the amount of available protein as well as activity per unit concentration. Thus, on both theoretical grounds and through numerous empirical examples^39-41^, MR analyses using variants in a gene encoding a drug target that affect its expression (or activity) have reproduced the effect direction of compounds with pharmacological action on the same protein^39-41^. Given the typically nonlinear drug dose-response, the small downstream effects of genetic variants on the level or function of a protein may underestimate the potential treatment effect of a drug. MR analyses assess the effect of target modulation in any tissue, whereas, certain tissues may be in accessible to a drug either because of its chemistry or the anatomical or physiological barriers. Furthermore, RCTs are closely monitored, and followed for a fixed period, allowing for exploration of induction-times^11^. MR estimates are considered to reflect a life-long exposure, but in the absence of serial assessment, possible changes across age are difficult to explore, as are disease induction-times. For these reasons we suggest that drug target MR offers a robust indication of effect direction but may not directly anticipate the effect magnitude of pharmacologically interfering with a protein. Findings such as the observed increased risk of AMD (CETP), asthma (PCSK9), Alzheimer’s in (PCSK9), therefore need to be considered in the context of both the duration of drug exposure and the potential for a drug to access the relevant tissues.

Our findings add to prior drug target MR analyses of CETP and PCSK9 which did not have access to genetic associations with protein concentration and weighted by downstream effects of the drug target on HDL-C or LDL-C, respectively. Here we showed consistency in the findings of MR analyses of CETP weighted through the concentration of the encoded protein (a more direct proxy of target modulation) and through HDL-C, TG, and LDL-C. Such “biomarker weighted” drug target MR should not be confused with MR analyses designed to evaluate the causal relevance of major lipid fractions; utilising genetic variants from throughout the genome^14^. In the presence of post-translation pleiotropy^14^, where perturbation of a protein affects multiple downstream biomarkers, some of which may lie on the causal pathway to disease and others not, biomarker weighted drug target MRs do not provide evidence on the possible mediating pathway of the drug target on disease^14^ and instead reflect drug target effects.

In conclusion, previous failures of CETP inhibitors are likely related to suboptimal target inhibition (dalcetrapib), off-target effects (torcetrapib) or insufficiently long follow-up (evacetrapib). The present drug target MR analysis, consistent with findings from the anacetrapib trials, anticipates that on-target CETP inhibition decreases CVD risk. MR analyses additionally suggests a reduction in kidney disease risk, but an increased risk of age-related macular degeneration.

## Data Availability

All data are publicly available, as described in the methods section. Please contact AFS for access to specific files, data, or analysis scripts.

## Author’s contributions

AFS, ADH, CF, contributed to the idea and design of the study. AFS and NH performed the systematic-review and meta-analysis. AFS, NH, and CF performed the analyses. AFS drafted the manuscript. All authors provided critical input on the analyses and the drafted manuscript.

## Guarantor

Amand F Schmidt performed the here presented analyses, had full access to all the data in the study and takes responsibility for the integrity of the data and the accuracy of the data analysis.

## Acknowledgement

This research has been conducted using the UK Biobank Resource under Application Number 12113. The authors are grateful to UK Biobank participants. UK Biobank was established by the Wellcome Trust medical charity, Medical Research Council, Department of Health, Scottish Government, and the Northwest Regional Development Agency. It has also had funding from the Welsh Assembly Government and the British Heart Foundation.

## Prior postings and presentations

The preliminary meta-analysis of RCT data were presented at BPS 2018 by NH. The preprint version of this manuscript has been deposited on medrxiv.

